# Analytic sensitivity of the Abbott BinaxNOW™ lateral flow immunochromatographic assay for the SARS-CoV-2 Omicron variant

**DOI:** 10.1101/2022.01.10.22269033

**Authors:** Sanjat Kanjilal, Sujata Chalise, Adnan Shami Shah, Chi-An Cheng, Yasmeen Senussi, Michael Springer, David R. Walt

## Abstract

The emergence of the SARS-CoV-2 Omicron variant has motivated a re-evaluation of the test characteristics for lateral flow immunochromatographic assays (LFIAs), commonly referred to as rapid antigen tests. To address this need, we evaluated the analytic sensitivity of one of the most widely used LFIAs in the US market, the Abbott BinaxNOW™ COVID-19 Ag At-Home Card using 32 samples of Omicron and 30 samples of the Delta variant. Samples were chosen to intentionally over-represent the range of viral loads where differences are most likely to appear. We found no changes in the analytic sensitivity of the BinaxNOW™ assay by variant even after controlling for variation in cycle threshold values in the two populations. Similar to prior studies, the sensitivity of the assay is highly dependent on the amount of virus present in the sample. While the analytic sensitivity of the BinaxNOW™ LFIA remains intact versus the Omicron variant, its clinical sensitivity is influenced by the interaction between viral replication, the dynamics of tissue tropism and the timing of sampling. Further research is necessary to optimally adapt current testing strategies to robustly detect early infection by the Omicron variant to prevent transmission.

## Introduction

The identification of infection by severe acute respiratory syndrome coronavirus-2 (SARS-CoV-2), the virus that causes coronavirus disease 2019 (COVID-19), aids the individual by prompting timely initiation of treatment and aids the public by allowing for separation of contagious hosts from susceptible secondary contacts. Lateral flow immunochromatographic (LFIAs) assays, commonly referred to as rapid antigen tests, are an important element of pandemic containment as they provide results within 15 minutes and can be performed by the public without supervision. However, a drawback is their lower analytic sensitivity relative to nucleic acid amplification based assays like the reverse transcriptase polymerase chain reaction (RT-PCR). In practice, the impact of this drop in sensitivity is limited to the brief window during which a person is efficiently transmitting live virus but still has viral loads below the limit of detection (LoD) of the LFIA. In prior waves of COVID-19, this time period is estimated to be days 1 to 2 post-infection^1^. Tests taken during that time window have a risk of being falsely negative and can lead individuals to transmit the virus to others unbeknownst to them.

The COVID-19 pandemic has spread in a series of waves due to the successive emergence of variants of concern (VOCs) that contain sets of mutations that confer selective advantages over prior lineages due to improved transmissibility, immune escape, and more efficient viral replication. These mutations are concentrated in the spike protein as it is the target of greatest immune pressure, but VOCs also contain single nucleotide polymorphisms in the nucleocapsid (N) protein, which encapsidates the viral RNA. The recently emerged Omicron variant has a mutation (P13L) and a deletion (d31-33) in the N-terminal domain, and two changes in the central linker domain (R203K and G204R^2^) that have not yet been fully characterized.

The N protein is the target analyte for the majority of LFIAs currently approved for emergency use by the US Food and Drug Administration (FDA). These assays typically use monoclonal antibodies coupled to nanoparticles to bind epitopes present on the SARS-CoV-2 N protein, which are then carried by capillary action to a detection line where the complex is bound by a second monoclonal antibody which binds elsewhere to the SARS-CoV-2 N protein. The resulting concentrated nanoparticle can be visualized by eye. The regions of the protein to which antibodies are raised are derived either from the wild type strain or early VOCs such as the Alpha variant. The precise epitopes targeted by the commercially-available LFIAs are proprietary, therefore independent evaluation is necessary to evaluate whether changes to nucleocapsid protein in the Omicron variant affect the analytic sensitivity of the test. Given the critical role that LFIAs play in early case detection and return to work assessments, quantifying changes in test performance is of high value to the scientific community, policy makers and the public.

In this study we report the analytic sensitivity of the widely used Abbott BinaxNOW™ COVID-19 Ag Card against the Omicron variant, using the Delta variant as a reference. Our primary hypothesis is that the analytic sensitivity of the Abbott BinaxNOW™ LFIA will be unaffected by the nucleocapsid mutations specific to the Omicron variant.

## Methods

We utilized 32 specimens positive for the Omicron variant and 30 positive for the Delta variant, collected between December 16th 2021 and December 18th 2021 as part of Harvard University’s asymptomatic SARS-CoV-2 student and staff screening program. During the time period of collection, >98% of the community was fully vaccinated. All samples were obtained from the anterior nares of individuals using a RHINOstic nasal swab (Rhinostics, Boston, MA) and initially placed into a sterile dry tube. Upon reaching the central testing laboratory, samples were eluted into 300μl of phosphate buffered saline and inactivated at 65°C for 30 minutes. All samples were received by the laboratory and eluted within 24 hours. Real-time RT-PCR was performed using the Quaeris SARS-CoV-2 assay with fluorescence and cycle thresholds (Cts) for the N and RdRP genes determined by the Applied Biosystems RT-PCR instrument running the QuantStudio 7 Design and Analysis Desktop Software, version 1.7. Details of the protocol can be found in the FDA Emergency Use Authorization summary for the Harvard University Clinical Laboratory (HUCL) assay^3^. The LoD of the assay is 2.5 RNA copies / μl.

Samples were genotyped using a multiplex PCR assay with primers specific to both variants. Based on the expected limit of detection (LoD) for the BinaxNOW™ LFIA^4^, we weighted our samples to over-represent those with Ct values below 30 as this is where differences in analytic sensitivity would most likely appear. The final analysis for the Delta variant samples consisted of 22 samples with Ct < 30 and 8 samples with Ct ≥ 30. The final analysis for the Omicron variant samples consisted of 24 samples with Ct < 30 and 8 samples with Ct ≥ 30.

After RT-PCR analysis, the specimens selected for this analysis were frozen once at -20°C. For LFIA testing, 50μl of each sample was placed in a 1.5ml microcentrifuge tube into which the kit-supplied swab was placed and rotated for 15 seconds. This volume was chosen based on a prior study that determined 50μl to be the optimal volume for balancing the ratio of analyte to kit-supplied running buffer for *in vitro* testing of samples with the BinaxNOW™^4^. Based on our observations, we noted that on average 5μl of sample remained in the tube after swab rotation. All subsequent steps for sample evaluation were per the assay’s Instructions For Use^5^. Briefly, six drops of kit-supplied running buffer were placed into the top hole of the card and swabs were inserted into the bottom hole and pushed upwards until the tip was in contact with the buffer. The cards were then sealed and read twice after 15 minutes by two independent readers blinded to the identity of the sample. A third reader was used for any discordant reads and a photo was taken of the card after the completion of the incubation period. All samples were run in triplicate.

Descriptive statistics were performed using chi-squared tests or Fisher’s exact test for categorical variables and 2-sample t-tests for comparing Ct values by variant. Logistic regression was used to estimate the odds ratio of a positive LFIA test given variant type after accounting for Ct value differences. All analyses were performed in R, version 4.1.2. This study was deemed non-human subjects research and approved by the Mass General Brigham Institutional Review Board (protocol 2021P003604).

## Results

A total of 62 samples were run in triplicate (30 Delta samples and 32 Omicron samples). Only 2 samples differed within technical replicates and there were no differences by reader, therefore all results represent the data obtained from a single replicate and reader 1. Table 1 shows the distribution of Ct values for the samples chosen for this analysis did not differ by variant.

**Table 1:** Baseline distribution of Ct values by SARS-CoV-2 variant. IQR, interquartile range.

| Variant | Sample size | Median Ct value (IQR) | p value |
| --- | --- | --- | --- |
| Delta | 30 | 27 (5.88) | 0.94 |
| Omicron | 32 | 27 (6.02) |  |

The BinaxNOW™ test was positive in 9 of 22 (41%) Delta samples and in 8 of 24 (33%) Omicron samples with Ct < 30. It was positive in 1 of 7 (12%) Delta samples and in 0 of 8 (0%) Omicron samples with Ct ≥ 30. There were no statistically significant differences between the variants with respect to test positivity for either Ct range (Table 2). The complete dataset for this study is available by request.

**Table 2:** Distribution of BinaxNOW™ LFIA results stratified by Ct value ranges and SARS-CoV-2 variant.

| Ct range | Variant | BinaxNOW™ result | N (percent) | p value |
| --- | --- | --- | --- | --- |
| <30 | Delta | Negative | 13 (59%) | 0.82 |
|  |  | Positive | 9 (41%) |  |
|  | Omicron | Negative | 16 (67%) |  |
|  |  | Positive | 8 (33%) |  |
| ≥30 | Delta | Negative | 7 (88%) | 1.00 |
|  |  | Positive | 1 (12%) |  |
|  | Omicron | Negative | 8 (100%) |  |
|  |  | Positive | 0 (0%) |  |

Figure 1 shows the distribution of positive and negative results by Ct value and variant. The expected LoD for the BinaxNOW™ is estimated to be 40,000 to 80,000 RNA copies per swab, which is derived from samples obtained prior to the emergence of the Delta variant^4^. This corresponds approximately to a Ct of 25 on the HUCL assay based on previously performed standard curve data normalized to the sample volume taken up by the swab. Two of 10 (20%) Delta variant samples and 5 of 11 (45%) Omicron samples were negative despite having levels of virus above the LoD.

**Figure 1:**
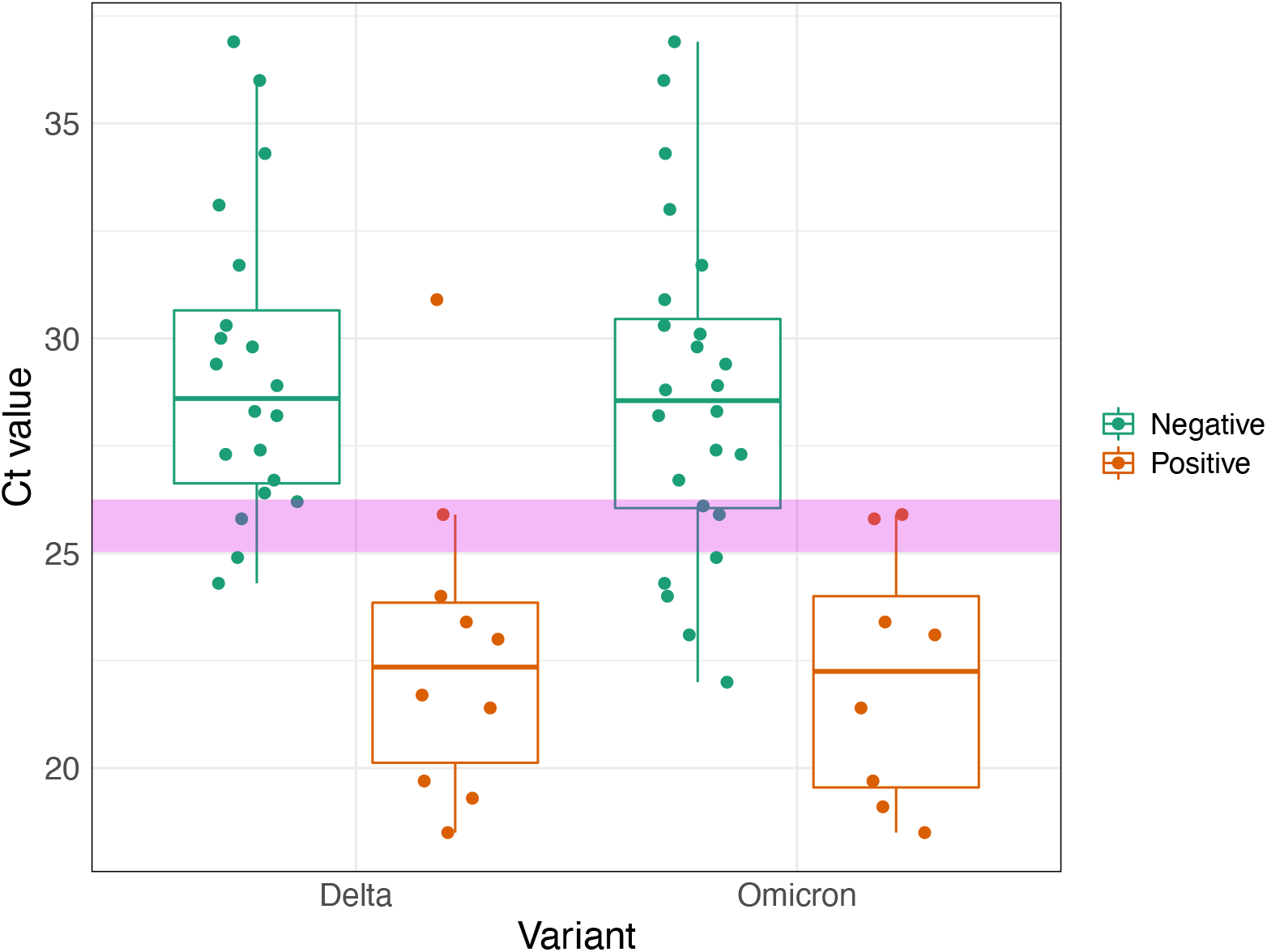
Distribution of BinaxNOW™ antigen test results by Ct value and SARS-CoV-2 variant. The y-axis indicates the Ct value of the positive sample. The pink horizontal band represents the expected LoD range of the BinaxNOW™, based on work from Perchetti et al^4^ and standard curve data for the HUCL RT-PCR assay.

After adjustment for each sample’s Ct value, there was no difference in the odds of test positivity for the Omicron variant versus the Delta variant (odds ratio 0.41; 95% CI 0.07 - 2.0).

## Discussion

The emergence of the Omicron variant in mid-November 2021 was followed by many anecdotal reports of false negative LFIA tests. In this study, we found no significant difference in the analytic sensitivity of the widely used Abbott BinaxNOW™ COVID-19 Ag Card LFIA for the N protein of the SARS-CoV-2 Omicron variant with respect to the Delta variant. We noted a greater proportion of negative samples containing viral loads above the assay’s LoD for Omicron specimens than for Delta specimens, but this finding did not reach statistical significance, possibly due to our study’s small sample size. Given that N protein mutations have been shown to impact LFIA performance in the past^6^, these findings provide a degree of reassurance that the assay is performing as expected.

Our results reinforce the findings of a smaller study that also examined the analytic sensitivity of the BinaxNOW™ versus Omicron with respect to its ability to detect both variants and in the estimated amount of nucleocapsid protein necessary to visualize a signal^7^. Our findings extend that work by including a larger number of independently collected clinical specimens and widening the range of viral loads in the sample set. Our results are also in agreement with the study by Deerain et al, which utilized serial dilutions of Omicron and Delta samples derived from viral cultures^8^. In that study, there was no difference in the analytic sensitivity for the Abbott Panbio™ COVID-19 Ag Rapid Test Device, which utilizes the same epitopes as the Abbott BinaxNOW™ COVID-19 Ag Card test marketed in the United States. Similarly, our findings agree with a recently published study from Schrom et al evaluating the analytic sensitivity of the BinaxNOW™ using paired RT-PCR anterior nares specimens obtained from a community testing center in San Francisco with a high rate of test positivity^9^. Our study provides complementary information to their findings by quantifying the threshold for detection in RNA copy numbers per swab and by comparing results with Omicron to Delta variant samples, which enables us to determine the degree of performance drift. Finally, our study is in line with announcements from the manufacturer^10^ regarding the performance of the LFIA, though the data underlying their press release are not publicly available.

Independent of whether the analytic sensitivity for the BinaxNOW™ remains intact, the real-world performance of the test may still substantially differ from its use during prior waves of the COVID-19 pandemic. This is because the clinical sensitivity of an assay is influenced by factors inherent to the virus as well as host factors such as pre-existing immunity. For instance, Abramson et al reported the propensity for the BinaxNOW™ and the Quidel Quickvue™ LFIAs to be falsely negative early in infection despite very high viral loads detected concomitantly from saliva samples^11^. Antigen detection from nasal swab samples eventually turned positive several days after symptom onset, suggesting that viral replication may initially localize to the oropharynx prior to transitioning to the nasopharynx. A study by Marais et al also found higher viral loads in saliva compared to paired mid-turbinate swabs but did not follow study participants over time to document a transition^12^. The BinaxNOW™ is currently only approved for use with anterior nares swabs in the US^5^.

Our results may be affected by the use of heat-inactivated samples that underwent one freeze-thaw cycle prior to analysis. However, this is not likely to have had a major impact on assay performance, as samples would have undergone denaturation regardless and the period of time they were frozen was short. The consistency of our data with prior studies defining the analytic sensitivity of the BinaxNOW™ as well as the lack of variation within technical replicates and across multiple blinded readers, further suggests that our findings are reproducible. Although our sample modality was not identical to a nares swab, we utilized a volume of media (50 μl) that has been shown to be approximately equal to the volume of mucus expected to be taken up by a swab when used *in vivo*^4^.

In summary, we found no difference in the analytic sensitivity of the Abbott BinaxNOW™ COVID-19 Ag Card for the Omicron variant relative to contemporaneously collected Delta variant samples. If sufficient virus is present in a sample, the assay should be expected to turn positive. The clinical (ie ‘real-world’) sensitivity of the assay is impacted by the viral load in the nares at the time of sampling, which in turn is influenced by variant-specific replication kinetics and host immunity. Increasing data suggests this may be the primary driver for the anecdotal reports of false negative LFIA results soon after symptom onset. Larger studies performing serial sampling from the oropharynx and nasopharynx with quantitative measures of viral loads in symptomatic and asymptomatic people will provide critical data for informing case detection algorithms.

## Data Availability

All data produced in the present study are available upon reasonable request to the authors.

## Acknowledgements

This study was funded by a grant from the Massachusetts Consortium for Pathogen Readiness.

